# Central and Brachial Pressures: Effects on Arterial Stiffness in Older Adults

**DOI:** 10.1101/2025.06.26.25330379

**Authors:** Sakar Gupta, Timothy Hess, Amy Hein, Claudia E Korcarz, Justyn Nguyen, Akinwale Iyeku, Jeremy R. Williams, Molly A. Cole, Ryan Pewowaruk, Adam D. Gepner

**Affiliations:** University of Wisconsin School of Medicine and Public Health, Madison, WI, USA; William S. Middleton Memorial Veterans Hospital, Madison, WI, USA

## Abstract

**Background:** Using brachial blood pressure (BP) as a surrogate for central BP when calculating carotid arterial stiffness (CAS) has not been studied in older adults.

**Methods:** Veterans (n=180) age 65+ were recruited from Madison VA Hospital. Resting supine brachial BP and central BP (estimated from radial artery waveforms, Atcor Medical) were obtained. CAS (Peterson’s elastic modulus [PEM], Young’s elastic modulus [YEM]) and distensibility coefficient (DC) were calculated using brachial and central BP. Differences in CAS were compared using paired Wilcoxon tests. Linear regression models evaluated associations with cardiovascular risk factors.

**Results:** Participants were 70.4 (7.7) years old and 27.8% were female. Average brachial systolic BP was significantly higher than central (132.3 [18.6] mmHg vs 123.8 [17.7] mmHg p<0.001). Compared to brachial BP, using central BP to calculate stiffness measures resulted in significantly lower YEM and PEM and significantly higher DC (PEM: 480.6 [209.5] mmHg vs 378.3 [178.4] mmHg; YEM: 2220.2 [926.6] mmHg vs 1746.9 [785.4] mmHg; DC: 2.4 [1.0] x10^−3^ mmHg^−1^ vs 3.1 [1.1] x10^−3^ mmHg^−1^; all p<0.001). Absence of hypertension was associated with smaller differences in PEM and DC (PEM: *β*=−29.1, SE=12.1, p=0.02; DC: *β*=−123.8, SE=55.3, p=0.027), while older age was associated with greater differences in YEM when calculated using brachial vs central BP (*β*=1.9×10^−5^, SE=0.69×10^−5^, p=0.006).

**Conclusions:** Brachial and central BP differ in older adults and result in significant differences in calculated CAS and distensibility. Brachial BP overestimates CAS, especially in those with hypertension.

## INTRODUCTION

Hypertension is characterized by the presence of persistently elevated blood pressure (BP), and it affects approximately 31.1% of adults globally.^1^ It is the leading modifiable risk factor for cardiovascular disease (CVD) and all-cause mortality worldwide.^1–4^ In clinical practice, BP is routinely assessed using cuff-based sphygmomanometry at the level of the brachial artery, a method that has long been the cornerstone for diagnosing and managing hypertension.

However, growing evidence suggests that central BP— reflecting pressures in the aorta and arteries proximal to the heart— is not always accurately represented by peripheral BP ^5–9^ Central BP more directly reflects the hemodynamic load experienced by end-organs including the heart, brain, and kidneys and may offer superior prognostic value for predicting cardiovascular health.^6–9^ This distinction becomes increasingly relevant with aging as the prevalence of hypertension rises, and peripheral BP has been shown to systematically overestimate central BP in older adults.^3,4,10^

The overestimation of peripheral BP is primarily due to pulse pressure magnification– a phenomenon in which the systolic component of the pulse wave increases as it travels away from the central aorta.^7,11^ Pulse pressure magnification is modulated by age-related changes in the arterial wall which ultimately lead to increased arterial stiffness and early wave reflections, and changes the gradient in vascular compliance across the arterial system.^7,11–13^

Arterial stiffness is also a key mediator of BP elevation and the progression to hypertension with aging.^14,15^ In older adults with hypertension, increased arterial stiffness is associated with adverse CVD outcomes.^16,17^ Importantly, most studies that derive local arterial stiffness measures use brachial BP measurements as a surrogate for central pressure.^13,18–21^ This assumption may introduce systematic error, particularly in older adults in whom pulse pressure amplification and arterial stiffness are more pronounced.

To address these knowledge gaps, our study evaluates the extent to which the discrepancy in using brachial BP compared to central pressures impacts the estimation of carotid arterial stiffness in at-risk, older Veterans. By comparing stiffness measures derived from both brachial and central BP, we aim to clarify whether current methodologies misrepresent vascular health in this population and whether central hemodynamic measurements should play a more prominent role in risk stratification and personalized hypertension management in older adults.

## METHODS

### Study Participants

Older (>60 years old), ambulatory, community dwelling Veteran participants (n=180) were recruited from the Madison Veterans Affairs Hospital and surrounding areas. The study was approved and reviewed by the University of Wisconsin Institutional Review Board and the Veterans Affairs Research and Development Committee. All participants gave written informed consent. Inclusion criteria were age >60 years old, able to read and write English, willing to undergo exercise treadmill stress testing administration of sublingual NTG, have a tonometric evaluation of arterial stiffness, and undergo a brachial and carotid artery ultrasound. Participants were excluded if they had a history of known CVD, secondary hypertension (aside from sleep apnea), chronic kidney disease (eGFR<30 mL/min/m2), active cancer, hypoxemic pulmonary disease, changing antihypertensive medication within 4 weeks of study participation, active rheumatologic diseases, human immunodeficiency virus, or any illness with an infectious etiology or fever >38°C or hospitalization for any reason within the prior 4 weeks. Each participant was initially identified as “normotensive” or “hypertensive” based on self-report or current anti-hypertensive medication use. Self-reported hypertensive participants were confirmed by evidence of two or more office BP readings with a systolic BP (SBP) > 130 mmHg or with a 24-hour ambulatory blood pressure monitor >125 mmHg.

### Study Protocol

All studies were conducted at the University of Wisconsin Atherosclerosis Imaging Research Program (UW AIRP) laboratory– a nationally recognized core ultrasound reading, training, and scanning laboratory.^22^ Before the initiation of the study, participants fasted for eight hours and were advised to abstain from sildenafil, smoking, and caffeine intake. Additionally, participants were directed to withhold their morning blood pressure medications on the day of their visit.

#### Brachial and Central BP measurements

Participants assumed a supine position in a temperature-controlled exam room and rested for 10 minutes. Serial baseline measurements of automated oscillometric brachial blood pressure were obtained (Cheetah Starling Fluid Management System, Baxter Healthcare, Deerfield, IL, USA). Central BP measurements were derived from radial artery tonometry wave forms (Atcor Sphygmacor, Atcor Medical, Sydney, Australia) using a Miller transducer and a validated and US Food and Drug Administration-approved transfer function.^23^

#### Carotid Arterial Stiffness (CAS) Calculations

Peterson’s Elastic Modulus (PEM), Young’s Elastic Modulus (YEM), and Distensibility Coefficient (DC) of the common carotid artery were calculated for each participant using both, the brachial and central BP measurements^23,24^:

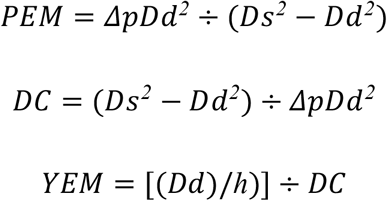

where Dd represents the internal arterial diameter at end-diastole, Ds represents the internal arterial diameter at peak systole, *Δp* represents the pulse pressure (PP), and h represents the intima-media wall thickness.

### Statistical Analysis

Peripheral and central BP measurements as well CAS measurements using both brachial and central values were compared using paired Wilcoxon tests (reported as average (St. deviation)). Linear regression models were used to evaluate associations between the difference in brachial and central BP/CAS measures and various CVD risk factors (reported as average [standard error]). The CVD risk factors that were analyzed included age, sex, hypertension status, diabetes, sleep apnea, alcohol use, active smoking status, and lifetime smoking status. A p-value < 0.05 was considered statistically significant for all comparisons.

## RESULTS

Baseline characteristics are described in Table 1. Participants were a mean (standard deviation) of 70.4 (7.7) years old and 72.2% (130) were men. The vast majority of participants were White 90.6% (163), 7.2% Black (13), and 1.7% (3) were Hispanic or Latino. 64.4% (116) had an existing diagnosis of hypertension, 18.9% (34) carried a diagnosis of diabetes, and 20% (36) had obstructive sleep apnea. 65% (117) of the participants reported actively consuming alcohol, averaging 5.4 (6.0) drinks per week, while 52.8% (96) disclosed having ever smoked tobacco, with 5.6% (10) reporting current active tobacco use.

**Table 1:** Baseline Characteristics of participants (n=180)

| Variable | All Participants (n=180) |
| --- | --- |
| Age (Years) | 70.4 ± 7.7 |
| Sex |  |
| Male (n, %) | 130 (72.2 %) |
| Female (n, %) | 50 (27.8 %) |
| Race |  |
| American Indian or Alaska Native (n, %) | 0 (0 %) |
| Asian (n, %) | 1 (0.6 %) |
| Black or African American (n, %) | 13 (7.2 %) |
| Native Hawaiian or Other Pacific Islander (n, %) | 0 (0 %) |
| White (n, %) | 163 (90.6 %) |
| Hispanic or Latino or Spanish Origin (n, %) | 3 (1.7 %) |
| Hypertensive Status (n, %) | 116 (64.4 %) |
| Diabetes Status (n, %) | 34 (18.9 %) |
| Obstructive Sleep Apnea (n, %) | 36 (20.0 %) |
| Alcohol Use (n, %) | 117 (65.0 %) |
| Average #Drinks/Week | 5.4 ± 6 |
| Smoking |  |
| Current (n, %) | 10 (5.6 %) |
| Lifetime (n, %) | 96 (52.8 %) |
| Blood Pressure Parameters |  |
| Brachial Systolic BP, mm Hg | 132.3 ± 18.6 |
| Central Systolic BP, mm Hg | 123.8 ± 17.7 |
| Brachial Diastolic BP, mm Hg | 79.2 ± 9.5 |
| Central Diastolic BP, mm Hg | 80 ± 9.6 |
| Brachial Pulse Pressure, mm Hg | 53 ± 16.4 |
| Central Pulse Pressure, mm Hg | 43.8 ± 15.3 |
| Arterial Stiffness Measures |  |
| Brachial Peterson's Elastic Modulus (mmHg) | 480.6 ± 209.5 |
| Central Peterson's Elastic Modulus (mmHg) | 378.3 ± 178.4 |
| Brachial Distensibility Coefficient (10<sup>-3</sup> mmHg<sup>-1</sup>) | 2.4 ± 1 |
| Central Distensibility Coefficient (10<sup>-3</sup> mmHg<sup>-1</sup>) | 3.1 ± 1.1 |
| Brachial Young's Elastic Modulus (mmHg) | 2220.2 ± 926.6 |
| Central Young's Elastic Modulus (mmHg) | 1746.9 ± 785.4 |

### Comparisons of Brachial vs Central BP and CAS measures

The average brachial systolic BP and PP were significantly higher than the central systolic BP (SBP: 132.3 mmHg (18.6) vs 123.8 mmHg (17.7); p<0.001; PP: 53 mmHg (16.4) vs 43.8 mmHg (15.3); p<0.001). When calculated using brachial BP measurements, PEM and YEM were significantly higher compared to when using central BP measurements (PEM: 480.6 mmHg (209.5) vs 378.3 mmHg (178.4); p<0.001; YEM: 2220.2 mmHg (926.6) vs 1746.9 mmHg (785.4); p<0.001). The DC was significantly lower when using brachial measurements compared to central BP (0.0024 mmHg^−1^ (0.001) vs 0.0031 mmHg^−1^ (0.0011), p<0.001). (Table 1, Figure 1).

**Figure 1.**
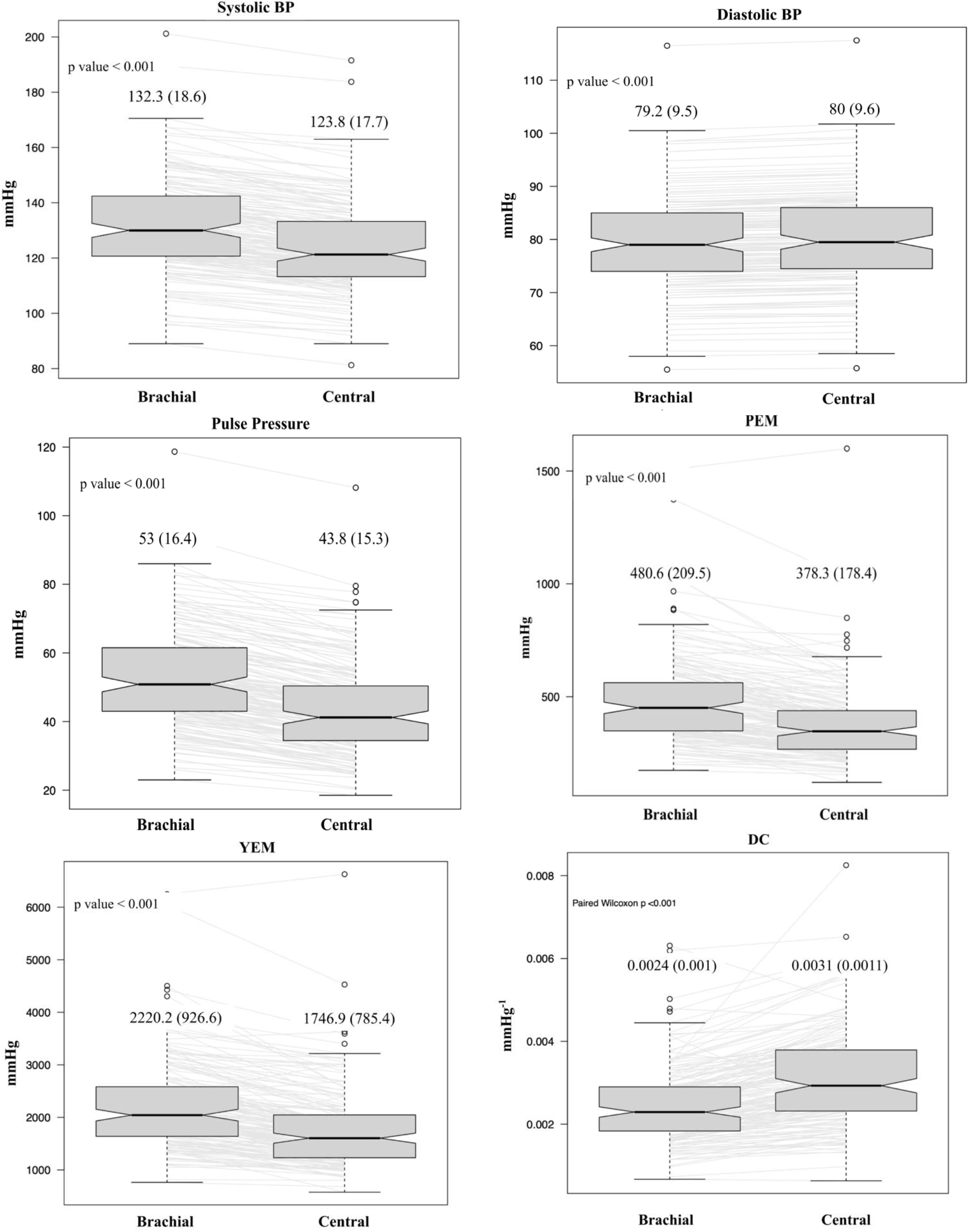
Comparisons of BP and CAS measures using Brachial and Central Pressures

### Linear Regression Models Evaluating Associations with CVD risk factors

Linear regression models evaluating associations between various CVD risk factors and the difference between brachial and central measurements are shown in Table 2. Smaller differences between brachial and central SBP and PP were strongly associated with younger age and female sex (SBP: β=−0.15, SE=0.042, p<0.001 and β=−0.17, SE=0.043, p<0.001 respectively; PP: β=−3.4, SE=0.73, p<0.001; β=−3.5, SE=0.74, p<0.001 respectively). The other CVD risk factors (hypertension status, diabetes, sleep apnea, alcohol use, active smoking status, and lifetime smoking status) did not show a strong association with differences between brachial and central BP measurements (Table 2). In terms of CAS measures, the absence of hypertension was strongly associated with smaller differences in brachial and central PEM (β=−29, SE=12, p=0.02) and DC (β=−123.8, SE=55.3, p-value=0.027). Furthermore, older age was associated with greater differences in brachial and central YEM (β=1.9×10^−5^, SE=0.69×10^−5^, p-value=0.006) (Table 2).

**Table 2:**
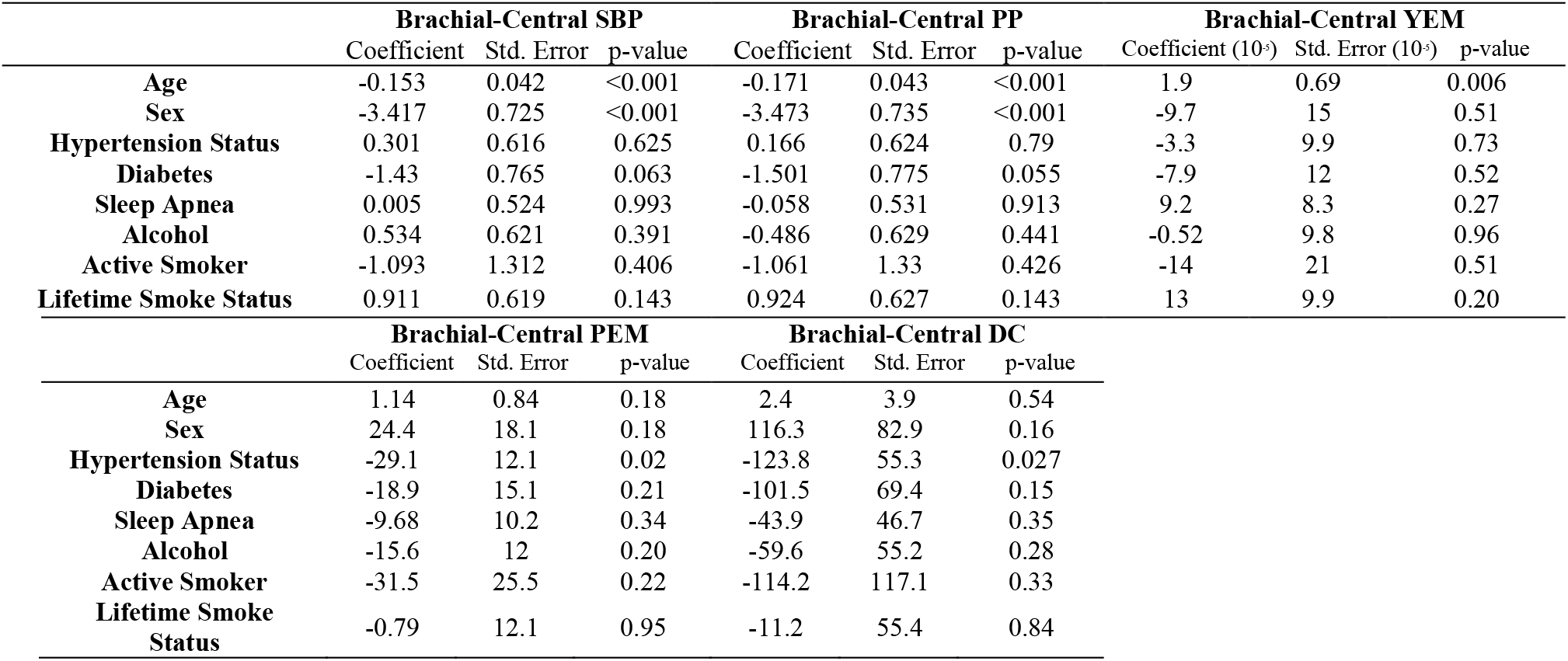
Linear regression models evaluating associations between differences in BP/CAS measures and CVD risk factors.

## Discussion

This analysis investigated the differences in central and brachial blood pressure measurements in older Veterans and the resulting impact on carotid arterial stiffness. Our study demonstrates that brachial SBP and PP are significantly higher than central SBP and PP in older adults. Furthermore, using brachial BP to calculate CAS significantly overestimates PEM and YEM and underestimates distensibility, especially in those with diagnosed hypertension.

The longitudinal effects of aging on CAS have been previously well described in a large multi-ethnic cohort over 10 years,^25^ however the CAS measures were calculated using brachial, not central blood pressures. In this cohort, on average, YEM of the carotid artery increased by 168 mmHg and DC decreased, on average, by *0. 4 x* 10^−*3*^*mmHg*^−*1*^ over 10 years indicating progression of arterial stiffness with aging. In a similar study of hypertensive adults, there was a 319 mmHg increase in YEM and *0*.*47 x* 10^−*3*^*mmHg*^−*1*^ decrease in DC on average over 10 years.^26^ Extrapolating the differences observed between central blood pressure and peripheral blood pressure seen in MESA to our analysis, calculating CAS using brachial BP overestimates YEM by 473.3 mmHg and underestimates DC by *0. 7 x*10^−*3*^*mmHg*^−*1*^. The difference in calculated local stiffness using brachial BP vs central BP correspond to approximately a decade’s worth of vascular aging in the older Veterans we studied.

Our regression analyses evaluated the influence of several CVD risk factors on the differences between brachial and central BP and CAS measures. Central and brachial SBP and PP measurements were the most different in older adults, where increased vascular stiffening with age and decreased arterial compliance likely contribute to greater pulse wave amplification. Interestingly, men had significantly greater discrepancies between brachial and central SBP and PP, possibly suggesting important differences in sex-based vascular characteristics. Other factors, such as diabetes, alcohol, smoking status, hypertension, and sleep apnea may play a less direct role in influencing brachial-central BP differences in this cohort. In terms of CAS, brachial and central PEM and DC were the most different in hypertensive participants and YEM was the most different in older adults, indicating that older patients with hypertension may experience the greatest divergence between brachial and central carotid artery stiffness. No significant associations were found for sleep apnea, sex, smoking status, diabetes, and alcohol use.

These findings emphasize that the differences in using central blood pressure measurements when calculating local CAS led to significant differences in arterial stiffness. This highlights the importance of incorporating central blood pressure into future studies that evaluate local arterial stiffness or consider alternative arterial stiffness measures that do not rely on BP (i.e. carotid to femoral pulse wave velocity), particularly in older, hypertensive adults. Based on our analysis, relying on brachial BP for CAS estimation may be reasonable in certain instances, particularly in younger females with normotension and without other vascular comorbidities where the difference between brachial and central BP is minimal. If calculations of CAS using peripheral blood pressures are being used for improved CVD risk prediction, the stiffness measures should be interpreted with caution, especially in older adults with hypertension.

### Limitations

While there are many strengths to this study, including recruiting an older adult population with and without hypertension, there are several limitations. The participants are community dwelling, ambulatory Veterans, who may be healthier than those who are non-ambulatory or institutionalized. Despite our attempts to over sample women and non-white Veterans, the majority of the participants were white, male veterans which limits the generalizability of the results to a broader population. Furthermore, our study did not assess lipid profile and renal function, which can impact blood pressure and arterial stiffness. Additionally, although participants were asked to hold their hypertension medications the morning prior to the study visits, the effects of longer acting medications could have persisted into the study period.^22^ Finally, while central pressures have improved accuracy, their clinical use is limited by financial constraints and the need for specialized equipment although newer validated brachial cuff oscillometric methods that estimate central pressures can provide a more accessible alternative.

## Conclusion

The findings from our study highlight the importance of considering central BP when evaluating CAS to predict cardiovascular health and better understand vascular aging, especially in older adults with hypertension. Relying solely on peripheral BP may lead to an overestimation of vascular risk in these populations. Future studies should prioritize using central BP measurements or CAS indices that are independent of BP to more accurately assess arterial health and improve CVD risk stratification. This can help better guide antihypertensive therapy, potentially improving outcomes and reducing the risk of over- or under-treatment.

## Data Availability

Data will be available upon request.

## Non-Standard Abbreviations and Acronyms

CAS: Carotid arterial stiffness
CVD: Cardiovascular disease
SBP: Systolic blood pressure
PP: Pulse pressure
PEM: Peterson’s elastic modulus
YEM: Young’s elastic modulus
DC: Distensibility coefficient

## ACKNOWLEDGEMENTS

We would like to thank the Veterans who volunteered to participate in this study. This material is the result of work supported with resources and the use of facilities at the Madison VA Hospital and the University of Wisconsin Atherosclerosis Imaging Research Program.

